# Examining the Efficacies of Different Machine Learning Algorithms on Predicting Future Potential Death from Heart Failure

**DOI:** 10.1101/2023.11.11.23298416

**Authors:** Osama Radi

## Abstract

1 in every 5 deaths is from heart failure. If this heart failure was able to be predicted, medical practitioners would be able to issue the proper preventative measures in order to prevent the fatal heart attack. As machine learning is gaining its place in medicine, one of the most commonly asked questions is which machine learning algorithms can be used where. This paper aims to find which machine learning algorithm is most efficacious in predicting future fatal heart disease. Two machine learning algorithms were evaluated in this study; namely linear regression models and k-nearest-neighbors models. The K-Nearest-Neighbors model was found to be most efficacious with an accuracy between 96.67% and 100% in predicting future heart failure. The reliability of this algorithm in predicting death from heart failure will surely prove useful in the future of treating at-risk patients.

## 1 Introduction

As machine learning is finding its place in medicine, one of the most commonly asked questions is which machine learning algorithms can be used where. There are many different machine learning algorithms, and they all have applications in different areas of medicine and physiology. The issue is that there is a large amount of overlap when it comes to which machine learning algorithms work in certain areas. It is not uncommon to see multiple algorithms being used to solve the same problem, even though there is often one that is more effective than the others. Along the same lines, heart failure lends itself very well to predictive machine learning algorithms; this is because there are many measurable factors that lead to heart failure. Although machine learning is an extremely powerful tool in predicting heart failure, much of the use of machine learning in this area of medicine is misguided. Due to the fact that multiple machine algorithms may work for predicting heart failure to a relatively high degree of accuracy, many machine learning algorithms are disregarded which may potentially have higher degrees of accuracy than the ones currently in use. Thus, research in the efficacies of different machine learning algorithms on predicting heart failure is necessary to determine which algorithms have the highest degrees of accuracy.

This study examines the efficacies of different machine learning algorithms on predicting potentially deadly instances of heart failure. Research has been done on various uses of machine learning algorithms on the area of heart failure; for example, Bobak J. Mortazavi et al. conducted research on various machine learning techniques to predict readmissions due to heart failure [2]. Additionally, Evanthia E. Tripoliti et al. created an algorithm which predicts adverse events caused by heart failure [4]. Also, Cameron R. Olsen used classification algorithms in attempt to predict heart failure [3]. Because of the lack of diversity in the use of machine learning algorithms specifically to predict potentially deadly heart failure, it is essential to examine a wide variety of machine learning algorithms to find which machine learning algorithms are most viable for the prediction of deadly heart failure.

The goal of this study is to find which machine learning algorithms work best for detecting potentially deadly heart failure. All machine learning algorithms in the study will be trained on the same data: that being clinical records for heart failure from the UCI machine learning repository [1]. This data records patient information and whether the patient had a “death event” (whether the patient experienced a fatal heart attack in the follow-up period). This study will look at which machine learning algorithm most accurately predicts whether a death event occurred based on the data provided to it.

## 2 Methodology

### 2.1 Preface

The data that all of the algorithms are trained upon comes from the clinical records mentioned above [1]. This data-set has 12 pieces of information on 299 patients (age, anaemia, creatinine phosphokinase, diabetes, ejection fraction, high blood pressure, platelets, serum creatinine, serum sodium, sex, smoking, time after information was taken, DEATH EVENT). 11 of the 12 pieces of information can be used as inputs, and whether the patient experienced a death event (fatal heart attack) is the output of all algorithms. Additionally, all algorithms were developed using the python module SciKit-Learn and all graphs present were created using matplotlib.

### 2.2 Linear Regression

Linear Regression is the simplest form of machine learning. A computer is given inputs and outputs, and thus attempts to create a line of best fit where the inputs match the outputs. Linear Regression has a large limitation which is especially prevalent in this case: linear regression returns values that are not binary. The data that the algorithm is trained upon has a binary value of either 0 for no death event or 1 for a death event. This limitation can be bypassed by rounding the outputs of the linear regression model to the nearest whole number, which is almost always between 0 and 1.

For linear regression, the model is most accurate when you exclude some of the possible inputs. The most accurate model that was found included only serum creatinine, ejection fraction, blood pressure, creatinine phosphokinase, and time after the medical information was taken.

Generally, in order for linear regression to work, there must be a linear relationship between the inputs and the outputs. In this case, there was no clear linear relationship between the inputs and whether the patient experienced a death event.

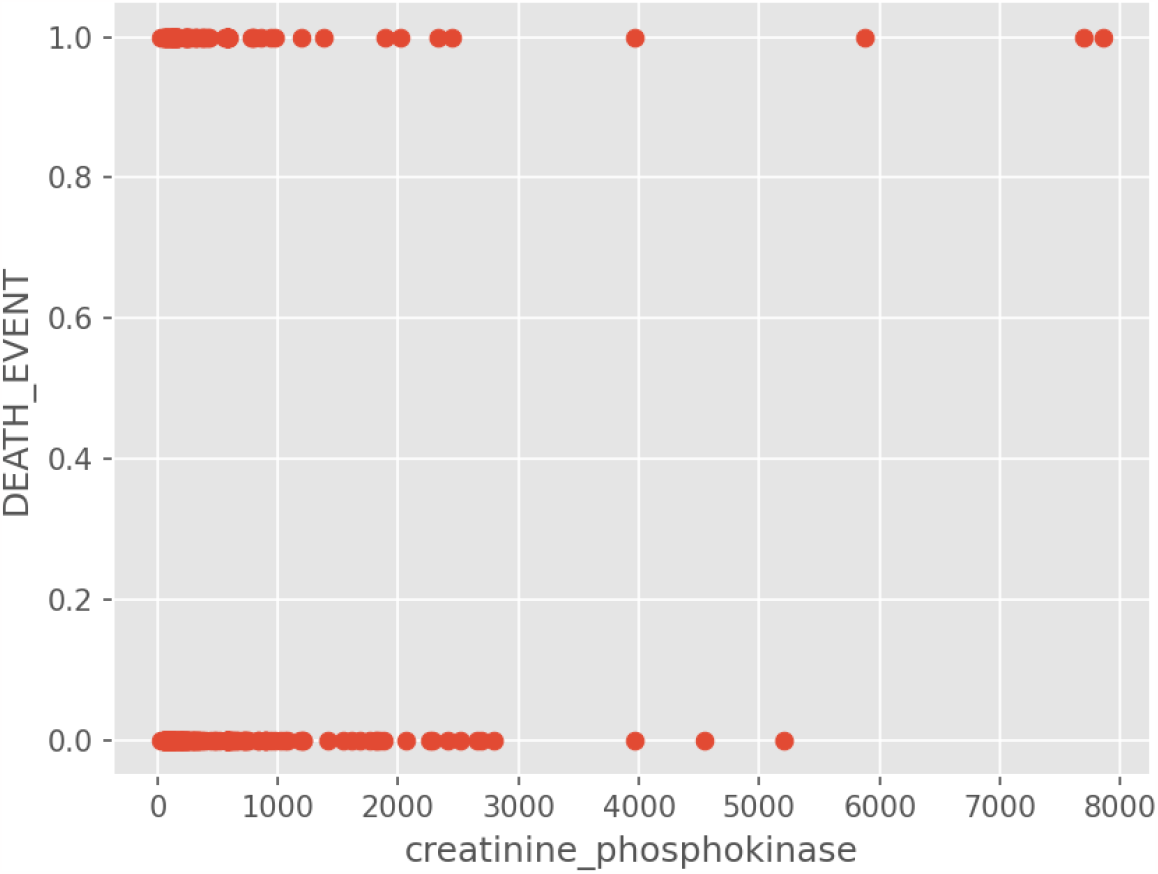

Relationship between creatinine phosphokinase and death event

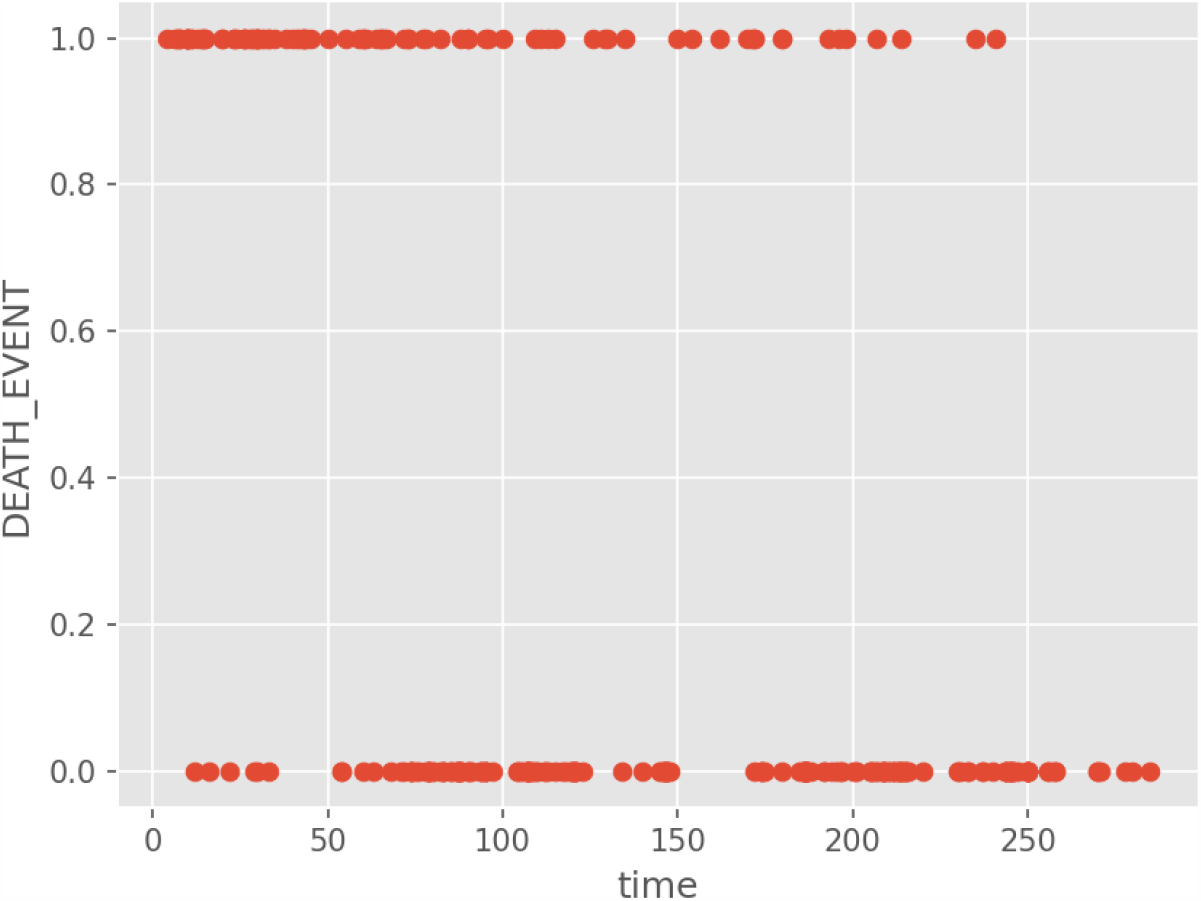

Relationship between time after data was taken and death event

### 2.3 K Nearest Neighbors

K Nearest Neighbors is a classification algorithm that evaluates a new data points class based on the classes of its nearest K (any arbitrary value) neighbors. This algorithm is extremely well suited for this case as it is classifying whether a patient will experience a death event. In this case, the patient data is inputted and it is put in to one of two classes: death event or no death event.

For the K Nearest Neighbors (KNN) algorithm, the accuracy of the predictions is highest when K (the number of neighbors evaluated) is 9. The predictions are also most accurates when only the following data is included: creatinine phosphokinase, age, sex, platelets, diabetes, ejection fraction, high blood pressure, serum creatinine, serum sodium, smoking, and time after information was taken.

For KNN to work most effectively, clusters of data points must mostly share the same class. In this case, there are examples of clusters sharing the same class.

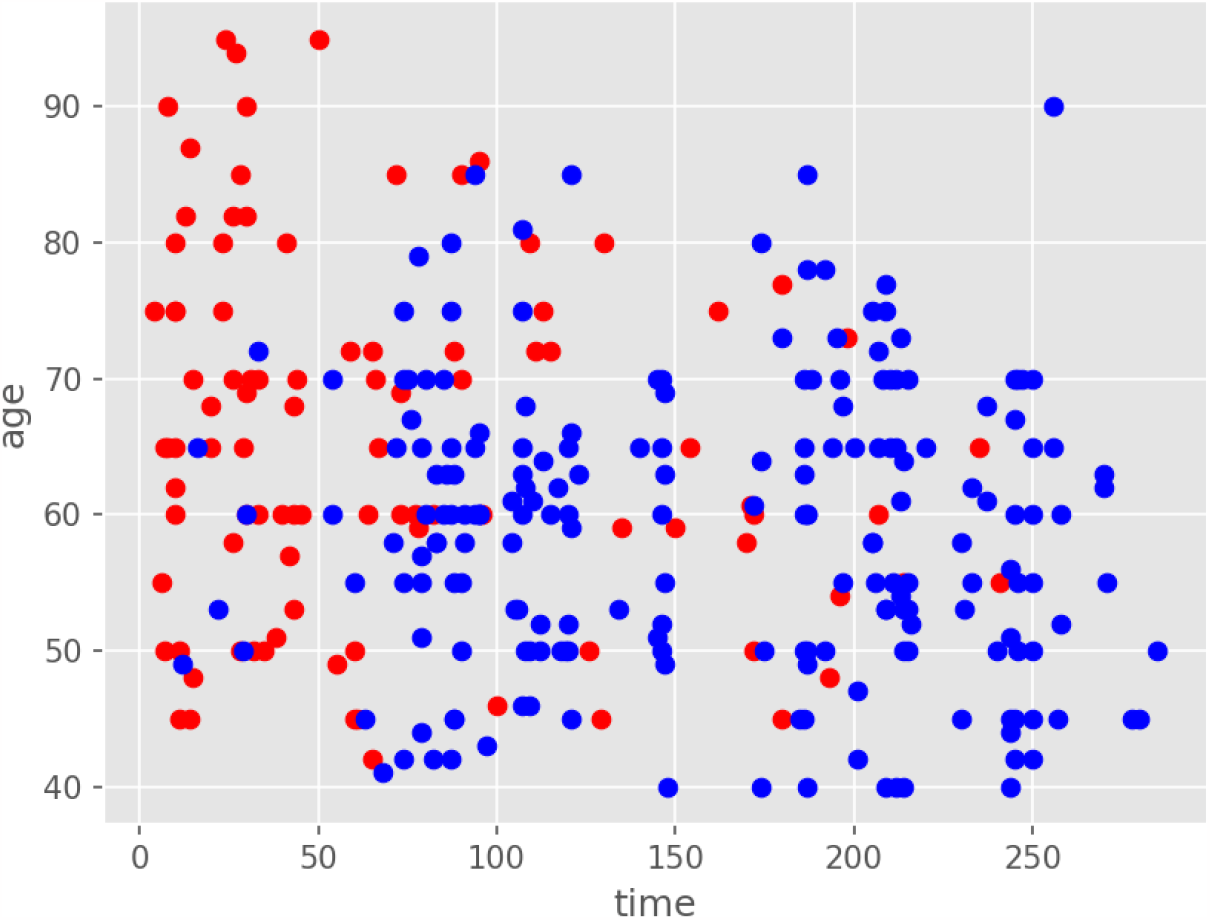

Relationship Between time and age (two arbitrary inputs). Blue = No Death Event, Red = Death Event

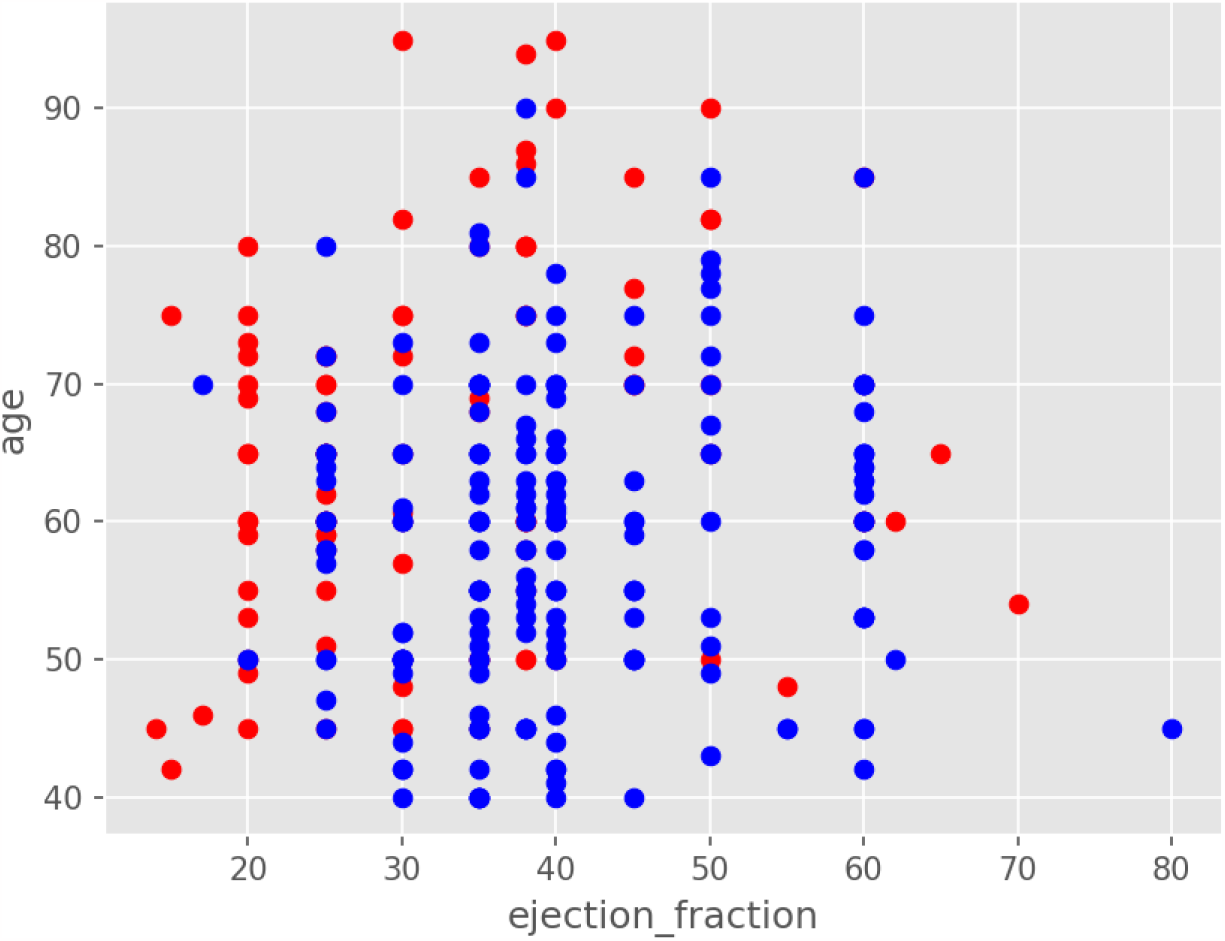

Relationship Between ejection fraction and age (two arbitrary inputs). Blue = No Death Event, Red = Death Event

You can very clearly see clusters of blue or red points on the graphs; thus, you can expect an accurate result.

## 3 Results

### 3.1 Preface

The accuracy of each algorithm was tested in the following manner. There were 299 patients total in the data set, 10% of the data was extracted at random as testing data, and the remaining 90% was used for training. The accuracy is determined by feeding the ≃ 30 patients’ data into the algorithms and comparing the real data to the algorithms’ predictions.

### 3.2 Linear Regression

Surprisingly, regardless of the lack of a clear linear relationship between the input data and the output data (Heart Failure), the linear regression model was able to have an accuracy score between 70.05% - 73.70%. The relatively high accuracy scores are likely due to the rounding effect mentioned in the methodologies section; this may have caused the algorithm to find a linear relationship when there wasn’t one that was clear.

**Example 1:** This is the raw data output of the linear regression model; a 0 represents no heart failure and a 1 represents heart failure.

| Actual | Predicted |  | Actual | Predicted |  | Actual | Predicted |
| --- | --- | --- | --- | --- | --- | --- | --- |
| 0 | 0 |  | 0 | 0 |  | 0 | 0 |
| 1 | 1 |  | 0 | 0 |  | 1 | 1 |
| 0 | 0 |  | 0 | 0 |  | 1 | 1 |
| 0 | 0 |  | 0 | 0 |  | 1 | 1 |
| 0 | 0 |  | 1 | 1 |  | 0 | 0 |
| 1 | 1 |  | 0 | 0 |  | 0 | 0 |
| 0 | 0 |  | 0 | 1 |  | 0 | 0 |
| 1 | 0 |  | 0 | 0 |  | 0 | 0 |
| 0 | 0 |  | 0 | 0 |  | 0 | 0 |
| 1 | 1 |  | 0 | 0 |  | 0 | 0 |

**Example 2:** This is the raw data output of the linear regression model; a 0 represents no heart failure and a 1 represents heart failure.

| Actual | Predicted |  | Actual | Predicted |  | Actual | Predicted |
| --- | --- | --- | --- | --- | --- | --- | --- |
| 0 | 0 |  | 1 | 1 |  | 1 | 1 |
| 1 | 1 |  | 0 | 0 |  | 0 | 0 |
| 0 | 0 |  | 0 | 0 |  | 0 | 0 |
| 0 | 0 |  | 0 | 0 |  | 0 | 0 |
| 0 | 0 |  | 0 | 0 |  | 1 | 1 |
| 0 | 0 |  | 0 | 0 |  | 0 | 0 |
| 0 | 0 |  | 0 | 0 |  | 0 | 0 |
| 0 | 0 |  | 1 | 1 |  | 0 | 0 |
| 0 | 0 |  | 1 | 1 |  | 1 | 1 |
| 1 | 1 |  | 0 | 0 |  | 0 | 0 |

### 3.3 K Nearest Neighbors

The K Nearest Neighbors model incredibly had an accuracy between 96.67% - 100%. In hindsight, this makes perfect sense, K-Nearest-Neighbors is a classification algorithm, but what is surprising is how accurate it is with the data provided; all of the data used can be collected from a short history and a blood test.

**Example 1:** This is the raw data output of the K Nearest Neighbors model; a 0 represents no heart failure and a 1 represents heart failure.

| Actual | Predicted |  | Actual | Predicted |  | Actual | Predicted |
| --- | --- | --- | --- | --- | --- | --- | --- |
| 0 | 0 |  | 1 | 1 |  | 0 | 0 |
| 0 | 0 |  | 1 | 1 |  | 0 | 0 |
| 1 | 1 |  | 0 | 0 |  | 0 | 0 |
| 0 | 0 |  | 0 | 0 |  | 1 | 1 |
| 1 | 1 |  | 0 | 0 |  | 1 | 1 |
| 0 | 0 |  | 0 | 0 |  | 0 | 0 |
| 0 | 0 |  | 1 | 1 |  | 1 | 1 |
| 0 | 0 |  | 0 | 0 |  | 0 | 0 |
| 0 | 0 |  | 1 | 1 |  | 0 | 0 |
| 0 | 0 |  | 1 | 1 |  | 0 | 0 |

**Example 2:**
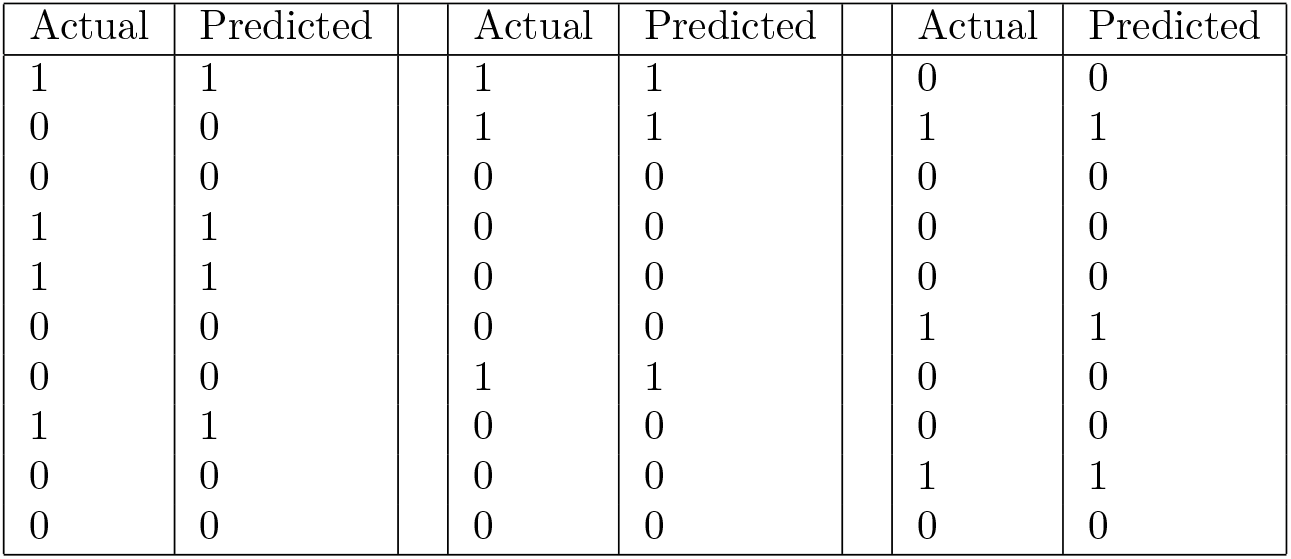
This is the raw data output of the K Nearest Neighbors model; a 0 represents no heart failure and a 1 represents heart failure.

| Actual | Predicted |  | Actual | Predicted |  | Actual | Predicted |
| --- | --- | --- | --- | --- | --- | --- | --- |
| 1 | 1 |  | 1 | 1 |  | 0 | 0 |
| 0 | 0 |  | 1 | 1 |  | 1 | 1 |
| 0 | 0 |  | 0 | 0 |  | 0 | 0 |
| 1 | 1 |  | 0 | 0 |  | 0 | 0 |
| 1 | 1 |  | 0 | 0 |  | 0 | 0 |
| 0 | 0 |  | 0 | 0 |  | 1 | 1 |
| 0 | 0 |  | 1 | 1 |  | 0 | 0 |
| 1 | 1 |  | 0 | 0 |  | 0 | 0 |
| 0 | 0 |  | 0 | 0 |  | 1 | 1 |
| 0 | 0 |  | 0 | 0 |  | 0 | 0 |

## 4 Discussion

The results of this study are extremely promising. The linear regression model unexpectedly could predict heart failure with an accuracy of over 70%, while the k-nearest-neighbors model could predict heart failure with an astonishing accuracy between 96.67% and 100%. These results definitively prove that the k-nearest-neighbors algorithm is the most efficacious algorithm to be used to predict fatal heart failure.

The results of this study definitively show the power that machine learning algorithms can have in a biomedical context. Using the k-nearest-neighbors algorithm, we can predict future heart failure with an astoundingly high accuracy; this can be used to issue the proper preventative measures before the heart failure occurs.

## 5 Conclusion

The aim of the study was to evaluate the efficacies of different machine learning algorithms based on their respective abilities to predict future heart failure. By evaluating the results, we were able to determine that k-nearest-neighbors is the most efficacious algorithm in this context as it has an accuracy which is between 96.67% and 100%.

These findings will be crucial in predicting heart failure before it is too late so that preventative measures can be issued. With this algorithm, medical practitioners will be able to predict future heart failure and hence issue the relevant medication and life style changes to prevent it before it is too late.

## Data Availability

All data produced in the present work are contained in the manuscript

https://archive.ics.uci.edu/dataset/519/heart+failure+clinical+records

## Notes

### Competing Interest Statement

The authors have declared no competing interest.

### Funding Statement

This study did not receive any funding

### Author Declarations

The data was collected from the UCI Machine Learning Repository. https://archive.ics.uci.edu/dataset/519/heart+failure+clinical+records

